# Assessing Cognitive Decline and Dementia Risk in Black and White Older Adults with Blood Biomarkers pTau217, GFAP, NfL, and Aβ ratio

**DOI:** 10.1101/2025.04.07.25325417

**Authors:** Ana W. Capuano, David A. Bennett, Jeffrey L. Dage, Kristen Russ, Konstantinos Arfanakis, Melissa Lamar, Lisa Barnes, Julie Schneider

## Abstract

**Background:** Alzheimer’s disease blood-based biomarkers are a cost-effective early-detection method that, for precision medicine reasons, needs to be studied in both Black and White. Few studies have a long follow-up of cognition. We studied a blood-biomarkers panel, cognitive decline, and dementia risk in a large number of Black and White participants.

**Methods:** Participants without dementia were followed annually up to 15 years, after plasma biomarker measurement (NfL, GFAP, amyloid-beta 42/40 ratio, pTau217). Data included demographics, medical history, blood tests (e.g., kidney function), MMSE, APOEε4, annual evaluations of cognition, and clinician-based dementia evaluation. The biomarkers’ association with comorbidities, cognitive decline, and risk of dementia was examined within race. To examine racial differences, we repeated analyses using a subset Mahalanobis-balanced 1:1 on sex, age, education, Latino/Non-Latino, and clinical status (hypertension, diabetes, GFR, BMI, and heart disease).

**Results:** Biomarkers were measured in 431 Black and 583 White (respectively mean age of 77 and 80, 17% and 21% men), generating a balanced sample of 366:366. Biomarker’s levels were similar between races. Within races, the associations of blood biomarkers with multiple comorbidities (especially kidney dysfunction and BMI) remained after controlling for demographics, APOE ε4, and dementia or death within 5 years. Men had lower GFAP levels than women (all p=<0.001). pTau217 was associated with decline in global cognition and all domains within races, and when comparing races, it was associated with a faster decline in global cognition and semantic memory in Black. The discrimination of dementia of pTau217 (AUC_3year, Black_ 0.81, CI=0.74, 0.89; AUC_3year, White_ 0.77, CI=0.71, 0.83) was good relative to age alone (AUC_3 year, Black_ 0.69, CI=0.58, 0.79; AUC_3 year, White_ 0.68, CI=0.62, 0.75), and MMSE (AUC_3 year, Black_ 0.81, CI=0.74, AUC_3 year, White_ 0.89; AUC_3 year, White_ 0.72, CI=0.65, 0.80). The discrimination of pTau217 did not improve by adding race or other biomarker information.

**Discussion:** pTau217 was highly associated with dementia risk and cognitive decline. The association of blood biomarkers with cognitive decline in Black and White participants was similar. Higher levels of pTau217 were, however, associated with semantic memory decline in Black adults. The combination of pTau217 with other biomarkers or MMSE did not improve dementia discrimination.

## INTRODUCTION

Dementia represents a growing public health challenge, with its prevalence rising alongside a population that is rapidly aging and becoming more diverse worldwide.^1^ Blood-based biomarkers, now recognized as less invasive and more accessible indicators of neurodegenerative processes, hold significant promise in pre-symptomatic diagnosis, enhancing diagnostic accuracy, and monitoring disease progression^2^.

Identifying biomarkers that can signal the onset and progression of dementia is crucial for early intervention and treatment strategies.

Among the emerging blood biomarkers, phosphorylated tau (pTau), neurofilament light chain (NfL), glial fibrillary acidic protein (GFAP), and the amyloid-beta 42/40 ratio (Aβ42/Aβ40) have garnered considerable attention for their potential roles in diagnosing and tracking Alzheimer’s disease and other dementias, including in diverse populations.^3, 4^. However, research on these biomarkers in Black older adults is relatively limited.^5–8^ Few have examined the blood biomarkers in relation to clinical measures and robust cognitive assessments. Studies evaluating the association between blood biomarkers and cognitive decline in older Black adults are also missing.

In our study, we examined a blood biomarker panel consisting of pTau217, NfL, GFAP, and Aβ42/Aβ40 among White and Black older adults to assess their associations with decline in global cognition and five cognitive domains. We also evaluated the blood biomarkers’ discrimination of risk of developing dementia alone in combination, and in relative to other clinical and functional assessments. Lastly, we conducted comparative analyses matching the sample of Black and White older adults, and this balanced sample to identify any differential patterns in biomarker levels and their relationship to cognitive outcomes, thereby contributing to the understanding of dementia risk factors across racial groups.

## METHODS

### Participants

Participants were from three longitudinal studies in the Rush Alzheimer’s Disease Center: the Religious Orders Study (ROS), the Minority Aging Research Study (MARS), and the African American Clinical Core (AACore)^9, 10^. ROS began enrollment in 1994 and includes older Catholic nuns, priests, and brothers, from across the US. MARS comprises Black older adults from the Chicago area and outlying suburbs and began recruitment in 2004. AACore, established in 2008, is focused on recruiting Black older adults from the Chicago area and outlying suburbs and primarily supports larger studies of Alzheimer’s disease and related dementias (ADRD) within and outside of the Rush Alzheimer’s Disease Center. Participants from all cohort studies enroll without known dementia and are followed annually until death. All have agreed to blood donation as part of their clinical evaluation. All cohorts use the same training, and testing staff, and include a large proportion of harmonized data that allows for merging the data across studies to increase power. All three cohort studies received IRB approval, and all participants signed consent and an Anatomic Gift Act. Data sharing is available under request at https://www.radc.rush.edu. Race and ethnicity are self-reported.

### Clinical and Cognitive Assessment

Each year, participants underwent a standardized clinical evaluation comprising a structured medical history, comprehensive neuropsychological testing, and a neurological examination. An experienced clinician subsequently classified dementia status based on the criteria established by the joint working group of the National Institute of Neurological and Communicative Disorders and Stroke (NINDS) and the Alzheimer’s Disease and Related Disorders Association (ADRDA). The diagnosis of dementia was made by a clinician with expertise in the field, following a review of cognitive testing results, neuropsychologist impressions, neurological exam data, and interview information. It necessitated evidence of cognitive decline and impairment in at least two cognitive domains, as previously outlined.^9,11–13^

Cognitive composites including global cognition and 5 cognitive domains were computed using 18 tests available for both cohorts, administered as part of each annual clinical evaluation^9, 14^. The global cognition composite was based on all 18 tests. The domains are based on subsets of tests including 7 measures of episodic memory (Word List Memory, Word List Recall, Word List Recognition; immediate and delayed recall of Logical Memory Story A and the East Boston Story), 3 measures of semantic memory (Boston Naming Test, Verbal Fluency, Word Reading), 3 measures of working memory (Digit Span Forward, Digit Span Backward, Digit Ordering), 3 measures of perceptual speed (Symbol Digit Modalities Test, Number Comparison, Stroop Test), and 2 measures of visuospatial ability (Standard Progressive Matrices, Judgment of Line Orientation). To calculate each composite, we converted raw test scores to z scores, using the baseline mean and SD in the combined cohorts, and averaged the z scores of component tests to obtain each composite score.

In addition, we collected the Mini-Mental State Examination (MMSE), a widely used 30-item standardized screening measure of cognitive function. In May 2021 the MMSE was replaced by the Montreal Cognitive Assessment (MoCA). MoCA items were used to ascertain a score equivalent to MMSE. The estimated MMSE score ranges from 0 to 30 and is either the MMSE score or the MMSE score based on the MoCA score.^15, 16^

Medical history of heart conditions and hypertension were self-reported. Participants were asked if they had ever been told by a doctor, nurse, or therapist that they had a heart attack or coronary, coronary thrombosis, coronary occlusion, myocardial infarction, or high blood pressure. History of stroke was based on clinician review of self-report questions, available neurological exam, cognitive testing, and interview of participant. History of claudication was obtained from self-reported pain in calves while walking. History of diabetes was obtained by self-reported diabetes, which included self-reported “sugar in the urine” or “high blood sugar” or self-reported medication use, including “insulin or injections for your high blood sugar” or “medicine by mouth for your high blood sugar”. In addition, a visual inspection of medications for diabetes was performed. Body mass index (BMI) was calculated by dividing weight (kilograms) by median height (meters) squared (kg/m2).

Glomerular filtration rate (GFR) was obtained to assess kidney function. A standard procedure was used to collect blood samples. Phlebotomists and nurses collected the blood specimen in a 2-mL EDTA tube using a sterile technique. Specimens were transferred to Quest Laboratories (Wood Dale, IL, USA) for a complete blood count analysis using a Beckan/Coulter LH750 automated processor. GFR using the 4-variable formula derived from the Modification of Diet in Renal Disease Study, where serum creatinine was determined using an Olympus AU4500 instrument at Quest Laboratories. Serum creatinine levels were not recalibrated to be traceable by isotope dilution mass spectrometry. In addition to the continuous GFR, we created a dichotomized measure of kidney function, such that measures equal to or above 60 mL/min/1.73 m2 were considered impaired kidney function and below were considered normal.

### APOE Genotyping

Blood was collected with acid-citrate dextrose anticoagulant, stored at room temperature, and processed for lymphocyte separation within 24 hours of collection. DNA was extracted from approximately 2 to 3 million cells using a Puregene DNA isolation kit (Gentra, Minneapolis, Minn), with APOE genotypes determined according to a method previously described.^17^ Genotyping was done blinded to all clinical data and dichotomized based on the presence of at least one copy of the APOE ε4 allele versus those without a copy.

### Blood Biomarkers for Alzheimer’s Disease

Plasma biomarker assays were run blinded in the National Centralized Repository for Alzheimer’s Disease and Related Dementias (NCRAD) Biomarker Assay Laboratory as previously described^18^. Laboratory quality system procedures are in place for initial assay qualification, maintaining the longitudinal performance of assays using monitoring when major changes occur (lot changes, instrument maintenance) and bridging when necessary. Major changes and bridging occur using a set of n>25 samples from our internal biorepository. Acceptance criteria are established for each assay and are consistent with the documented performance of each assay. Every run contains pooled plasma reference samples which are used to control chart and monitor performance across, plates, days, instruments, operators, and studies. Overall study quality performance measures are available in the supplemental data file. Each assay plate (31 samples) was balanced on age, gender, and collection visit for longitudinal samples. Plasma samples were centrifuged at 8,324xg for 5 minutes before analysis.

Each sample was processed in duplicate per the manufacturer’s instructions starting with 4x or 3x dilution (N4PE and pTau217, respectively) with kit-specific diluent on a Tecan Fluent 1080 automated liquid handler. Plasma neurofilament light chain (NfL), glial fibrillary acidic protein (GFAP), beta-amyloid 1-42 (Aβ42), beta-amyloid 1-40 (Aβ40), and phosphorylated tau at threonine 217 (pTau217) were analyzed utilizing the Quanterix Simoa Neurology 4-Plex E Advantage Kit and the ALZpath pTau217 assay on a Quanterix Simoa HD-X. All assays were performed according to the manufacturer’s instructions except for the centrifugation speed due to cryovial sizes. Manufacturer-provided quality control (QC) samples or endogenous quality controls (eQCs) were used to verify results met quality criteria for analytical performance provided by the manufacturer as well as lab-specific expectations (Supplementary Material).

### Statistics

Initial analyses include examining the distribution of the panel of blood biomarkers, and the association of the biomarker panel with demographics and comorbidities within race. Amyloid-beta is treated as the ratio of 42 over 40 (Aβ42/Aβ40). Biomarkers in the original scale were mostly not normally distributed, and we used median, interquartile range, and non-parametric tests such as the U-test in initial analyses. Next, we applied a log2 transformation on the biomarkers in simple linear regression with the biomarkers as an outcome. These models included terms for multiple comorbidities and adjusted for demographics (age, sex, and education), dementia within 5 years, death within 5 years, and the presence of APOE ε4 allele. The comorbidities studied included heart disease, diabetes, kidney dysfunction, and BMI.

The association of the biomarker panel with the decline in cognition within race was assessed using linear mixed effect models with terms for standardized biomarkers from the original scale and adjusting for age biomarker measurement, years of formal education, sex, BMI, impaired kidney function, history of heart conditions, history of hypertension, phone interview indicator, and their interaction with time from study entry. To predict the time-specific risk of dementia of each biomarker we used the time-dependent-receiver operation curve (ROC) estimated using Inverse Probability of Censoring Weighting (IPCW) to evaluate the performance of the biomarker panel.^19^ Time-dependent AUC was ascertained using the control defined as a participant that is not a case^20^, and a 95% confidence interval was computed using Bootstrap. To provide a clinical perspective on the discriminant ability of the blood-based biomarkers we also calculated the discrimination of age alone and blood-based clinical assessments including the MMSE and, secondarily, RADaR (Rapid Assessment of Dementia Risk).^21^

Next for a rigorous comparison between races, we identified a subset of Black and White participants with similar profiles. For that, we used a Malahanobis matching algorithm with age within 5 years and education within 4 years by indicators of sex, ethnicity (Latino/Non-Latino), having at least 2 longitudinal evaluations, clinical status (presence of hypertension, diabetes, high GFR, underweight or BMI <18.5, history of congestive heart failure and heart disease). In this matching process, we could identify a balanced group of 366 White and 366 Black older adults. This balanced sample of 732 individuals was used for overall racial comparison purposes. Note that we only have 4 Latinos so we don’t perform ethnicity comparisons here. Among these, 716 had at least 2 longitudinal observations and were used in analyses of change in cognition. Analyses were performed in SAS/STAT software, Version 9.4 of the SAS® system for Linux, and R version 4.4.1 (Survival and Risk Regression packages).

## RESULTS

Plasma of 1014 older adults (431 Black and 583 White participants) without dementia at baseline were tested for blood biomarker measures (Table 1), followed for a median of 5 years. We examined the influence of multiple comorbidities (history of heart disease, hypertension, diabetes, and impaired kidney function) on the blood biomarker levels within race. We used a linear model with the log of each blood biomarker as the outcome and terms for the multiple morbidities controlling for demographics (age, sex, and education), APOE ε4 allele status, dementia within 5 years, and death within 5 years (Supplementary Table). Impaired kidney function was associated with pTau217, NfL, GFAP (all p<=0.011), and Aβ42/Aβ40 in Black participants. BMI was inversely associated with NfL, and GFAP (all p<001) in both races, and with pTau217 in Black participants. In this model, we also identified a strong sex difference for GFAP (all p<0.001) with lower levels in men of both races. APOE ε4 allele status remained associated with higher levels of pTau217 in White participants only.

**Table 1.** Demographics of Black older adults and the balanced sample with White older adults.

|  | Total sample n=1014 |  |  |  | Balanced Sample n=732 |  |  |  |
| --- | --- | --- | --- | --- | --- | --- | --- | --- |
|  | Blacks<br>n= 431 |  | White<br>N=583 |  | Blacks<br>n= 366 |  | Whites<br>n= 366 |  |
| Demographics |  |  |  |  |  |  |  |  |
| Age, mean (std) | 76.68 | (6.56) | 80.62 | (7.29) | 77.10 | (6.68) | 79.07 | (6.34) |
| Formal education in years,<br>mean (std) | 15.24 | (3.17) | 18.86 | (2.99) | 15.63 | (3.05) | 17.96 | (2.38) |
| Men, n (%) | 72 | (17%) | 122 | (21%) | 63 | (17%) | 63 | (17%) |
| APOE ε4 allele, n (%) | 136 | (37%) | 107 | (22%) | 113 | (37%) | 67 | (22%) |
| Clinical variables |  |  |  |  |  |  |  |  |
| Body mass index, mean (std) | 30.63 | (6.57) | 28.27 | (5.98) | 30.10 | (6.20) | 28.50 | (6.28) |
| Memory complaint, n (%) | 128 | (30%) | 162 | (28%) | 105 | (29%) | 96 | (26%) |
| History of heart conditions*, n<br>(%) | 36 | (8%) | 64 | (11%) | 27 | (7%) | 43 | (12%) |
| History of hypertension*, n<br>(%) | 343 | (80%) | 351 | (61%) | 284 | (78%) | 237 | (65%) |
| History of diabetes**, n (%) | 131 | (30%) | 112 | (19%) | 97 | (27%) | 85 | (23%) |
| History of stroke*, n (%) | 38 | (9%) | 50 | (9%) | 30 | (8%) | 27 | (8%) |
| History of claudication*, n<br>(%) | 32 | (22%) | 135 | (23%) | 29 | (21%) | 87 | (24%) |
| GFR, mean (std) | 72.77 | (18.06) | 75.93 | (15.20) | 73.35 | (18.16) | 74.63 | (16.47) |
| Impaired kidney function†, n<br>(%) | 117 | (28%) | 115 | (20%) | 89 | (25%) | 88 | (25%) |
| Brain function |  |  |  |  |  |  |  |  |
| MMSE, median (IQR) | 28 | (27-29) | 29 | (28-30) | 28 | (27-29) | 29 | (28-30) |
| Global Cognition, mean (std) | -0.03 | (0.55) | 0.37 | (0.54) | -0.01 | (0.55) | 0.42 | (0.53) |
| Blood Biomarker |  |  |  |  |  |  |  |  |
| pTau217, median (IQR) | 0.42 | (0.31,0.64) | 0.54 | (0.28,0.86) | 0.42 | (0.31,0.65) | 0.56 | (0.37,0.82) |
| NfL, median (IQR) | 17.43 | (12.49,26.74) | 22.35 | (15.91, 30.96) | 14.42 | (12.73,26.74) | 22.01 | (18.91,30.13) |
| GFAP, median (IQR) | 152.80 | (104.65,222.03) | 182.23 | (126.47,246.64) | 155.10 | (110.46,225.48) | 181.48 | (123.74,246.48) |
| Aβ42/Aβ40, median (IQR) | 0.05 | (0.05,0.06) | 0.05 | (0.04,0.06) | 0.05 | (0.05,0.06) | 0.05 | (0.04,0.06) |

\*self-reported \*\* self-reported and medication ‡ based on a GFR equal to or above 60 mL/min/1.73m<sup>2</sup>
GFR=glomerular filtration rate, MMSE= Mini-Mental State Examination

### Cognitive decline and dementia discrimination within race

To examine the association of the biomarker panel with cognitive decline within race we ran mixed-effect models fully adjustedfor demographics and comorbidities. A total of 412 Black and 544 White participants had 2 or more measures of global cognition and were included in these models. Table 2 shows the association of the standardized biomarkers with change in cognition over time, estimated using linear adjusted mixed-effect models with cognition as the outcome. pTau217 was associated with a decline in global cognition and all five cognitive domains in both races (all p<.0001). GFAP was associated with a decline in global cognition and all cognitive domains except visuospatial processing in both races, although the evidence of association with episodic memory in White participants was weak (β=-0.019, p=0.058). All blood biomarkers were associated with working memory decline, except Aβ42/40 in both races. Note that the direction of the association of Aβ42/Aβ40 was different from the other biomarkers as seen previously.^22^

**Table 2.** The effect of blood biomarkers on the change in cognition of Black older adults using linear mixed effect models.

| Interaction term with time | Estimate (Standard Error, p-value) |  |  |  |  |  |
| --- | --- | --- | --- | --- | --- | --- |
|  | Global cognition | Episodic Memory | Semantic Memory | Working Memory | Perceptual Speed | Visuospatial Memory |
| Black, n=412 |  |  |  |  |  |  |
| pTau217 | **-0.081(0.009,<.0001)** | **-0.083(0.011,<.0001)** | **-0.121(0.012,<.0001)** | **-0.056(0.01,<.0001)** | **-0.048(0.01,<.0001)** | **-0.041(0.011, <.001)** |
| NfL | **-0.012(0.005,0.023)** | **-0.013(0.006,0.035)** | -0.012(0.008,0.122) | **-0.022(0.006, <.001)** | -0.008(0.005,0.108) | 0.006(0.006,0.314) |
| GFAP | **-0.027(0.007, <.001)** | **-0.036(0.008,<.0001)** | **-0.039(0.01, <.001)** | **-0.019(0.008,0.014)** | **-0.028(0.008, <.001)** | -0.012(0.008,0.164) |
| Aβ42/Aβ40 | 0.01(0.005,0.071) | 0.01(0.006,0.134) | 0.014(0.007,0.058) | 0.001(0.005,0.864) | 0.001(0.006,0.82) | 0.006(0.006,0.312) |
| White, n=544 |  |  |  |  |  |  |
| pTau217 | **-0.051(0.007,<.0001)** | **-0.057(0.009,<.0001)** | **-0.064(0.009,<.0001)** | **-0.046(0.007,<.0001)** | **-0.058(0.007,<.0001)** | **-0.044(0.008,<.0001)** |
| NfL | -0.019(0.015,0.193) | 0(0.018,0.988) | -0.014(0.02,0.487) | **-0.063(0.017,<.001)** | **-0.044(0.016,0.006)** | -0.016(0.018,0.375) |
| GFAP | **-0.027(0.008,0.001)** | -0.019(0.01,0.058) | **-0.031(0.01,0.003)** | **-0.027(0.007, <.001)** | **-0.019(0.008,0.02)** | -0.005(0.009,0.594) |
| Aβ42/Aβ40 | 0.012(0.006,0.063) | **0.017(0.008,0.027)** | 0.015(0.008,0.063) | **0.011(0.006,0.048)** | **0.013(0.006,0.028)** | 0.001(0.006,0.912) |

In separate models stratified by race. Models with terms for standardized biomarkers in the original scale and their interaction with time from biomarker measurement, adjusting for age biomarker measurement, years of formal education, sex, body mass index, impaired kidney function, history of heart conditions, history of hypertension, phone interview indicator, and their interaction with time. GFAP =Glial Fibrillary Acidic Protein, NfL =Neurofilament Light Chain, Aβ42= beta-amyloid 1-42, Aβ40= beta-amyloid 1-40, and pTau217=phosphorylated tau at threonine 217.

Next, we examined the time-specific risk of dementia of each biomarker within race. In the total sample, 71 Black (16%) and 140 White (24%) older adults developed dementia, and 84 Black (19%) and 151 White (26%) older adults died before developing dementia. Based on the Reverse Kaplan-Meier^23^, Black participants had a median follow-up of 8 years (IQR 4 to 11 years), and White participants had a median follow-up of 7 years (IQR 4 to 14 years). In line with the previous studies of clinically relevant assessments^21^, we first examine a 3 to 7-year prediction. For this timeframe, the discrimination of biomarkers was good for Black, and White older adults, except for Aβ42/40 which performed poorly for both groups (Supplementary Figure). In particular, the discrimination of pTau217 was very good with a respective AUC_3year_ of 0.81 (95% CI=0.74, 0.89) and 0.77 (95% CI=0.71, 0.83) for Black and White older adults. Figure 3 provides the ROC curves of the biomarkers at 3 years.

**Figure 1.**
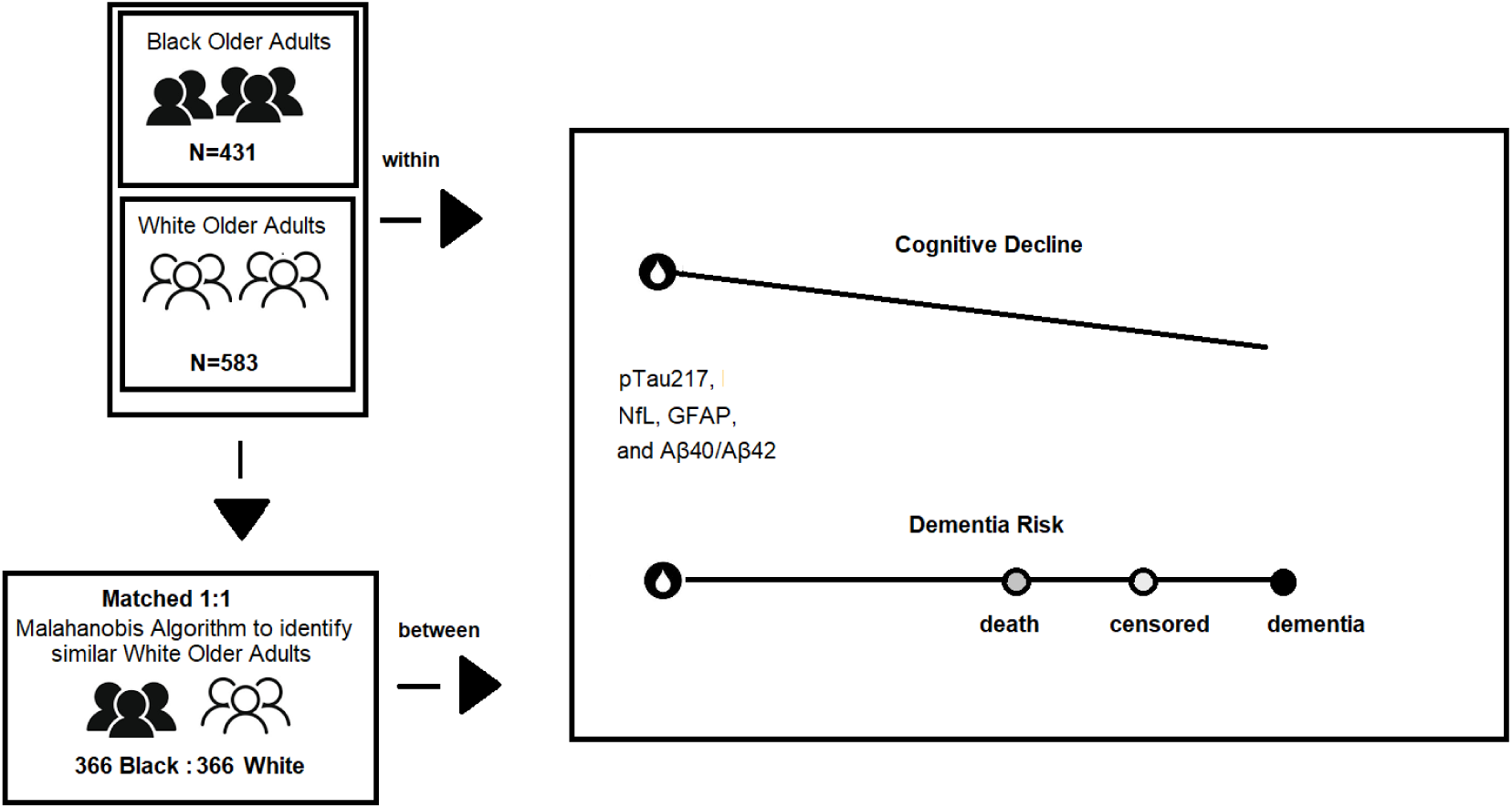
The schematic of the study design and association with key variables

**Figure 2.**
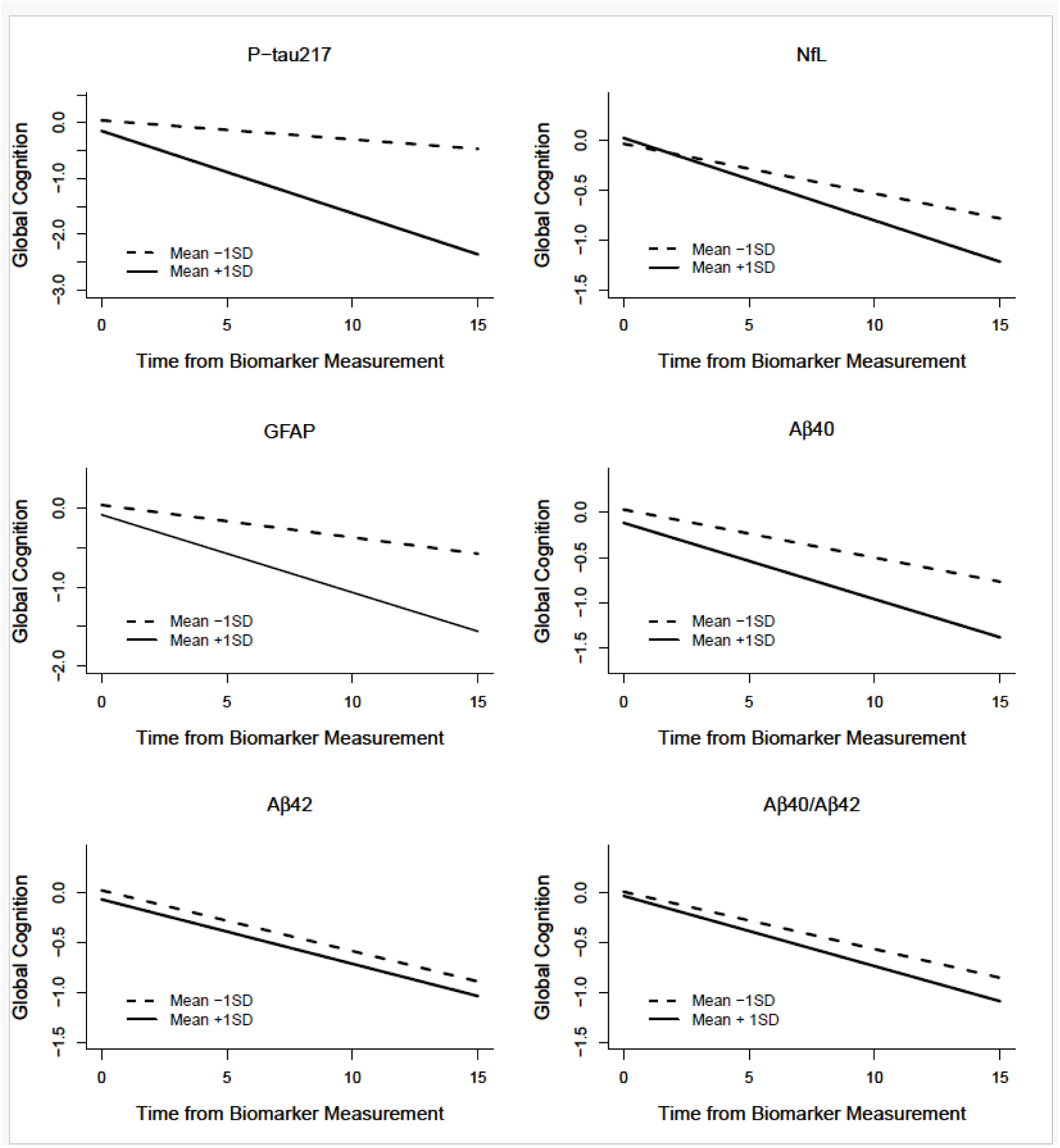
The effect of blood biomarkers on the trajectories of global cognition among Black older adults using adjusted linear mixed effect models.

**Figure 3.**
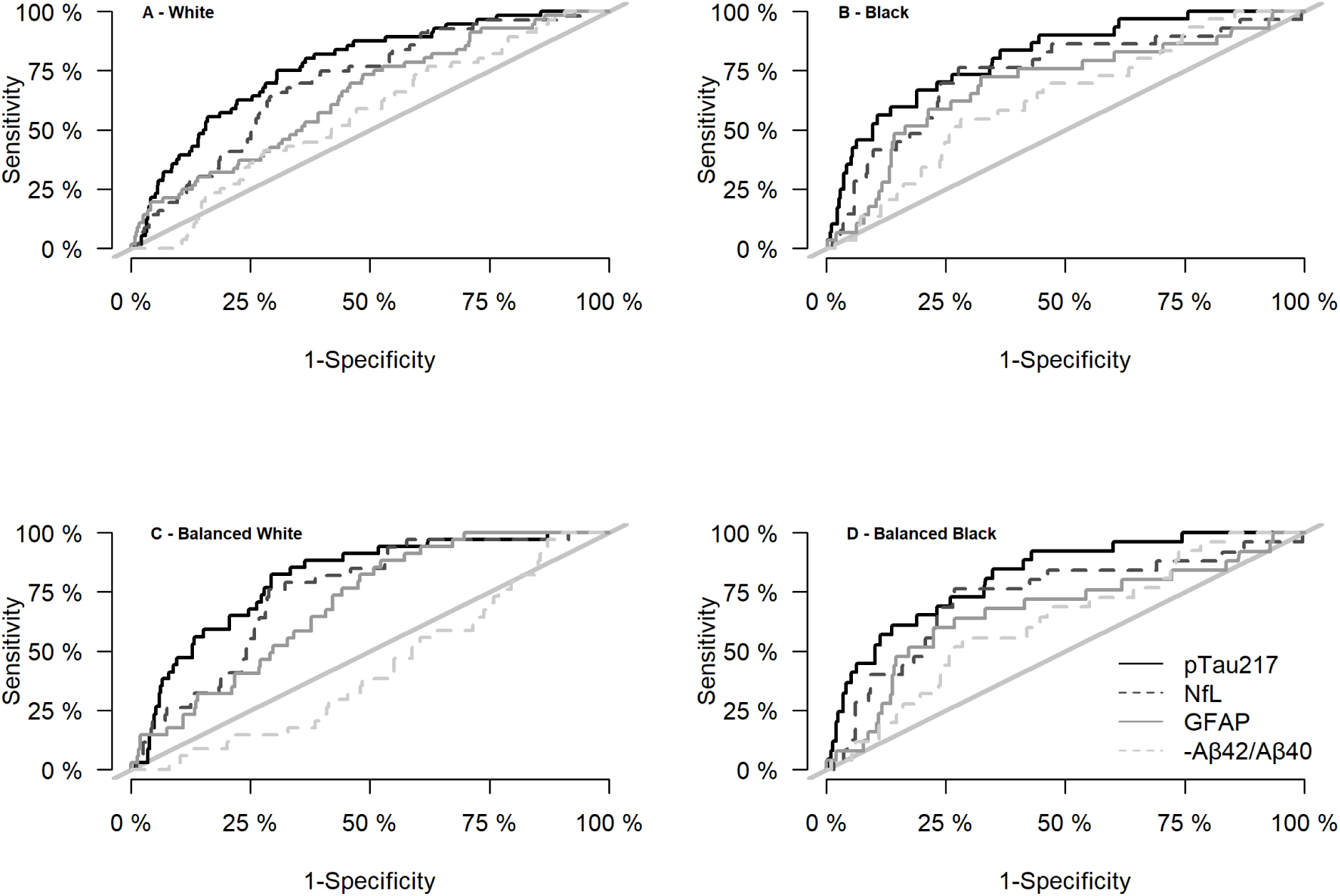
ROC curves of blood biomarkers at 3-year prediction time to dementia. Graphs A and B include the total sample of White and Black older adults. Graphs C and D include the balanced sample of White and Black older adults. GFAP =Glial Fibrillary Acidic Protein, NfL =Neurofilament Light Chain, Aβ42= beta-amyloid 1-42, Aβ40= beta-amyloid 1-40, and pTau217=phosphorylated tau at threonine 217.

To assess the biomarker’s utility in the clinical setting, we contrasted it with age alone and MMSE. Age alone had an AUC_3 year_ of 0.69 (CI=0.58, 0.79) and 0.68 (CI=0.62, 0.75), and MMSE had an AUC_3 year_ of 0.81 (CI=0.74, 0.89) and 0.72 (CI=0.65, 0.80), respectively in Black and White individuals. We next tested the discrimination of using multiple biomarkers combined or in combination with pTau217. The discrimination of using both PTau217 and the NfL was 0.81 (CI= 0.73, 0.89) for Black, and 0.70 (CI=0.59, 0.81) for White, representing a nominal loss compared to PTau217 alone, although again not enough to reach significance in this sample. Secondarily we compare the biomarkers with a recently developed quick risk score validated in Black older adults (RADaR, results in supplementary material).

In the final analyses, we estimated the false positive rate (FPR), true positive rate (TPR), positive predictive value (PPV), and negative predictive value (NPV) of dementia at 3 years considering death as a competing risk both for the total and balanced samples. A Black individual with pTau217 in the upper quartile (0.63) had an NPV of 67.9-69.9% and a PPV of 47.7-48.2%. A White individual with the same pTau217 had an NPV of 76.1.9-76.6% and a PPV of 48.5-50.0%.

### Racial comparisons

To ensure a rigorous comparison between races, we also identified a balanced subsample of Black and White participants (n=732, 366 per group) with similar age, education, ethnicity, clinical status, comorbidities, and longitudinal data availability. The description of the balanced groups is available in Table 1. Overall, most of the crude median values of the biomarkers were smaller for Black individuals. pTau217 had a median of 0.42 (IQR=0.31, 0.65) in Black and 0.52 (IQR=0.38, 0.82) in White individuals. Nfl had a median of 17.42 (IQR=12.7, 26.7) in Black and 22.01 (IQR=15.9, 30.1) in White individuals.

GFAP had a median of 155.1 (IQR=110.5, 225.1) in Black and 181.48 (IQR=124.1, 246.2) in White individuals. Aβ42/Aβ40 had a median of 18.9 (IQR=16.5, 21.4) in Black and 20.1 (IQR=17.6, 22.8) in White individuals.

Next, we ran a saturated linear regression model adjusting for demographics (age, sex, and education), dementia within 5 years, death within 5 years, the presence of APOE ε4 allele, and individual comorbidities (heart disease, diabetes, kidney dysfunction, and BMI). No racial differences were identified except for weak evidence of a difference in levels of pTau217 (p=0.054), which was lower in Black older adults. Next, we repeated the models, retaining BMI and impaired kidney function and adding the interaction of the comorbidities with race. There was no differential effect by race on the association of BMI or impaired kidney function with the biomarkers.

To compare Black to White older adults on the relation of the biomarkers to cognitive decline, we used 318 Black and 318 White individuals of the balanced sample who had 2 or more measures of global cognition. Table 3 shows the race effect on the association of each biomarker on the change of cognitive function, estimated using linear adjusted mixed-effect models with cognition as the outcome. pTau217 was differentially associated with a decline in global cognition, as well as semantic and visuospatial processing, depending on race. There was also weak evidence of a differential effect of NfL with working memory decline. Because APOE effects on cognition are known to be different in Black individuals, as a sensitivity analysis, we repeated these models using 200 Black and 200 White individuals from the balanced sample who did not have an APOE ε4 allele. In these analyses, the effect size of the differential association with global cognition was the same but it was no longer significant, likely because of power. The differential association with semantic memory decline was retained and a differential association with working memory decline was revealed.

**Table 3.** The differential effect of blood biomarkers on the change in cognitive function based on race in a balanced sample of Black and White older adults.

| Interaction term with race and time | Estimate (Standard Error, p-value) |  |  |  |  |  |
| --- | --- | --- | --- | --- | --- | --- |
|  | Global cognition | Episodic Memory | Semantic Memory | Working Memory | Perceptual Speed | Visuospatial Memory |
| Balanced sample, n=716 |  |  |  |  |  |  |
| pTau217 | **-0.03(0.012,0.018)** | -0.017(0.015,0.263) | **-0.068(0.016,<0.001)** | -0.018(0.012,0.133) | 0.000(0.014,0.979) | -0.025(0.013,0.053) |
| NfL | 0.009(0.015,0.551) | -0.002(0.018,0.929) | 0.000(0.022,0.987) | 0.023(0.018,0.214) | **0.031(0.016,0.045)** | 0.004(0.018,0.804) |
| GFAP | 0.006(0.011,0.548) | -0.007(0.013,0.61) | -0.013(0.014,0.38) | 0.009(0.011,0.379) | -0.007(0.011,0.507) | -0.013(0.011,0.23) |
| Aβ42/Aβ40 | -0.001(0.01,0.903) | -0.003(0.011,0.764) | 0.006(0.012,0.658) | -0.006(0.008,0.482) | -0.006(0.009,0.531) | 0.012(0.008,0.146) |
| Excluding APOE ε4 allele, n=400 |  |  |  |  |  |  |
| pTau217 | -0.031(0.016,0.057) | -0.031(0.022,0.166) | **-0.067(0.02,0.001)** | **-0.052(0.017,0.002)** | -0.012(0.018,0.51) | -0.026(0.02,0.18) |
| NfL | -0.021(0.034,0.526) | 0.016(0.045,0.73) | -0.041(0.044,0.344) | -0.065(0.035,0.067) | 0.044(0.036,0.224) | 0.01(0.04,0.802) |
| GFAP | -0.004(0.013,0.761) | -0.024(0.017,0.158) | -0.013(0.017,0.439) | -0.015(0.013,0.267) | -0.006(0.014,0.675) | -0.021(0.015,0.155) |
| Aβ42/Aβ40 | 0(0.013,0.988) | 0(0.016,0.999) | 0.013(0.016,0.416) | -0.009(0.012,0.433) | 0.003(0.012,0.804) | 0.023(0.013,0.071) |

Model with terms for standardized biomarkers in the original scale, race and their interaction with time from biomarker measurement, adjusting for age biomarker measurement, years of formal education, sex, body mass index, impaired kidney function, history of heart conditions, history of hypertension, phone interview indicator, and their interaction with time. GFAP =Glial Fibrillary Acidic Protein, NfL =Neurofilament Light Chain, Aβ42= beta-amyloid 1-42, Aβ40= beta-amyloid 1-40, and pTau217=phosphorylated tau at threonine 217.

Finally, to understand if race would improve discrimination, we repeated the analyses using the balanced sample with both PTau217 and race. The resulting AUC_3year_ was 0.78 (95% CI=0.69, 0.86), which did not represent an important gain in discrimination.

## Discussion

In this study involving 431 Black and 583 White older adults, we measured blood biomarkers of pTau217, NfL, GFAP, and Aβ42/Aβ40 and followed participants annually for a median of 5 years. We first examined each racial group separately using a larger sample and carefully adjusting for demographics and comorbidities as appropriate. Next, we conducted rigorous racial comparisons using a subset of Black and White participants, who were carefully matched on demographics, comorbidities, and other factors. There was, overall, a strong association between impaired kidney function and higher levels of pTau217, NfL, GFAP, and between lower levels of BMI and higher levels of NfL and GFAP. Within Black individuals, impaired kidney function was also associated with higher levels of Aβ42/Aβ40, and lower BMI was associated with higher pTau217. There was also a sex difference in levels of GFAP in both Black and White adults, such that men had lower values. Other studies with younger participants have also found lower levels of GFAP in men^24^. Regarding cognitive decline, higher levels of pTau217 were associated with faster decline in both races but consistently associated with faster decline in semantic memory in Black compared to White participants. GFAP was associated with a decline in global cognition and all cognitive domains except visuospatial processing in Black participants. There was no racial difference in the association between GFAP and cognitive decline. Finally, regarding the ability to predict dementia, among biomarkers, pTau217 had very good discrimination. In particular for Black participants, the AUC_3year_ of pTau217 was 0.81 (95% CI=0.74, 0.89), the same AUC_3 year_ of MMSE (0.81, CI=0.74, 0.89). However, combining pTau217 with other blood tests or clinical assessments did not improve the discrimination ability. These results suggest that pTau217 is a robust biomarker for cognitive decline and dementia risk, independent of other biomarkers and common risk metrics in both Black and White adults. Notably, the superior discrimination of pTau217 to the risk of dementia aligns with previous studies, suggesting the potential for greater generalizability of these biomarkers across diverse cohorts, thereby enhancing their clinical applicability.

Recent studies have indicated a significant association between pTau217 levels and various comorbidities^6, 7, 25, 26^, including cardiovascular disease, diabetes, kidney disease, hypertension, and BMI, as well as differential effects of APOE ε4 allele by race. However, few of these studies include a large number of Black older adults and many don’t adequately adjust findings to verify if these associations are not due to other factors. Although we found an unadjusted association among several comorbidities and the biomarkers, low BMI and impaired kidney function remained associated with higher levels of many biomarkers after adjustments. One could postulate that these comorbidities are associated with biomarkers because they are also a risk for dementia. One study highlighted the nuanced relationship between plasma phosphorylated tau levels and several comorbidities, suggesting that conditions such as cardiovascular disease and impaired kidney function not only contribute to a higher risk of dementia but may also interact with amyloid pathology^26^. In our work, however, we controlled for conversion to dementia and, separately, death in five years, and the associations remained. More studies should be conducted to further understand these associations, potentially including brain amyloid pathology.

Most AD biomarker studies have been conducted with White older adults. However, there is a slowly growing literature examining these biomarkers among older Black adults and findings have been mixed. For example, some studies of Aβ ratio demonstrate that amyloid-beta ratios are on average higher^27–29^, or lower^30^ between Black and White older adults. Some studies show no adjusted difference^4, 29, 31^ in pTau (total, pTau181, pTau231 or pTau217) between races. There are fewer studies of racial differences in NfL or GFAP, but here as well findings have been mixed, with some studies showing higher levels of NfL^32^, lower^27^ or equal^29^ among Black compared to White adults. Some studies have shown differences in plasma biomarkers^4, 6, 33^. Reasons for the mixed results are unclear but likely result from differences in participant recruitment (clinic vs community-based), adjustment for comorbidities (e.g., kidney function or BMI), or confounding by demographic features. When we adjusted our sample of Black and White adults on key demographic factors, most of the racial differences were eliminated.

Even fewer studies have linked biomarkers to cognitive test performance or dementia status in Black adults, and to our knowledge, no study has examined the relation of biomarkers to the annual rate of cognitive decline. Two studies^4, 6^ examined pTau181, pTau217, total tau (t-tau), and amyloid-beta (Aβ) levels in Black individuals. They found that among Black participants, elevated levels of p-tau181 and pTau217 were significantly linked to poorer cognitive performance. Using our balanced sample with further adjustments, we find in our study a race difference in the association of pTau217 biomarker with faster semantic memorydecline. It would be important to replicate these differences using other cohorts to see if this difference holds.

Our study has several strengths and few weaknesses. It studieda large number of Black older adults, to expand the understanding of how to utilize these tests in this population. Black and White individuals underwent similar annual evaluations by design and had blood biomarkers collected from the same laboratory. Black individuals experience a higher prevalence of comorbidities such as impaired kidney function, so another strength of the study was the ability to consider multiple comorbidities. A limitation, however, is that these are on average educated, healthy volunteers, many with a religious

lifestyle. The racial differences observed in the relationship between pTau217 and domain-specific cognitive performance highlight the necessity for tailored research. With varying findings in the literature regarding biomarkers such as NfL and GFAP, our study also reinforces the need for larger, longitudinal investigations that account for demographic and health-related differences, ultimately guiding targeted interventions.

## Data Availability

Data sharing is available under request at https://www.radc.rush.edu.

## Acknowledgments

We thank the MARS, ROS, and AA Clinical Core participants, the Rush Alzheimer’s Disease Center team (among several Annie Barz, Dominika Burba, John Gibbons, Ryan Johnson, Shahram Oveisgharan, Theresa M Mathes, Tiffany Wu, Traci Colvin, Wenqing Fan, Woojeong Bang), NCRAD’s biomarker assay laboratory (Jacob Wohlgamuth).

This work was funded by the National Institute on Aging, and the National Institute of Neurological Disorders and Stroke grants: R01AG022018, P30AG10161, P30AG072975, R01AG15819, and U24 AG021886.

ROS, MARS, and AACore resources can be requested at www.radc.rush.edu.

---

Supplementary table: saturated models with multiple comorbidities, genetics, and 5-year conversion to dementia or death and biomarkers as outcome. Linear regression models ran within each race. Biomarkers were log2 transformed in these models.

|  | pTau217 | NfL | GFAP | Aβ42/Aβ40 |
| --- | --- | --- | --- | --- |
| **Black** |  |  |  |  |
| Age in years centered at 78 | 0.019 (0.006), 0.004 | 0.033 (0.007), <0.001 | 0.035 (0.006), <0.001 | -0.007 (0.003), 0.052 |
| Men | 0.017 (0.101), 0.864 | -0.055 (0.114), 0.631 | **-0.385 (0.1), <0.001** | 0.04 (0.052), 0.444 |
| Years of formal education centered at 17 | 0.015 (0.012), 0.205 | 0.006 (0.014), 0.674 | 0.002 (0.012), 0.858 | 0.002 (0.006), 0.777 |
| APOE ε4 allele status | 0.103 (0.079), 0.191 | -0.005 (0.089), 0.955 | 0.039 (0.078), 0.619 | -0.031 (0.041), 0.444 |
| Body mass index | **-0.018 (0.006), 0.004** | **-0.027 (0.007), <0.001** | **-0.026 (0.006), <0.001** | 0.002 (0.003), 0.562 |
| Impaired kidney function | **0.394 (0.088), <0.001** | **0.664 (0.098), <0.001** | **0.298 (0.086), 0.001** | **0.117 (0.045), 0.01** |
| History of heart conditions | 0.085 (0.142), 0.55 | 0.161 (0.16), 0.316 | 0.111 (0.14), 0.43 | 0.038 (0.073), 0.606 |
| History of diabetes or medication | 0.103 (0.086), 0.234 | 0.156 (0.097), 0.11 | -0.002 (0.085), 0.98 | -0.023 (0.045), 0.603 |
| History of hypertension | 0.098 (0.099), 0.325 | 0.062 (0.112), 0.579 | 0.007 (0.098), 0.945 | -0.119 (0.051), 0.021 |
| Dementia in 5 years | 0.696 (0.131), <0.001 | 0.459 (0.147), 0.002 | 0.313 (0.13), 0.016 | -0.069 (0.068), 0.311 |
| Died in 5 years | 0.277 (0.113), 0.014 | 0.802 (0.127), <0.001 | 0.15 (0.111), 0.178 | 0.016 (0.058), 0.787 |
| **White** |  |  |  |  |
| Age in years centered at 78 | 0.013 (0.007), 0.08 | 0.029 (0.006), <0.001 | 0.021 (0.005), <0.001 | -0.005 (0.003), 0.113 |
| Men | 0.104 (0.109), 0.341 | -0.038 (0.085), 0.655 | **-0.347 (0.083), <0.001** | 0.007 (0.052), 0.886 |
| Years of formal education centered at 17 | -0.007 (0.014), 0.594 | -0.018 (0.011), 0.086 | -0.01 (0.01), 0.323 | 0.004 (0.006), 0.526 |
| APOE ε4 allele status | **0.384 (0.096), <0.001** | 0.027 (0.074), 0.717 | 0.016 (0.073), 0.823 | -0.08 (0.046), 0.08 |
| Body mass index | -0.01 (0.008), 0.211 | **-0.024 (0.006), <0.001** | **-0.03 (0.006), <0.001** | 0.006 (0.004), 0.087 |
| Impaired kidney function | **0.34 (0.113), 0.003** | **0.579 (0.087), <0.001** | **0.219 (0.086), 0.011** | 0.09 (0.053), 0.094 |
| History of heart conditions | -0.209 (0.134), 0.122 | **0.267 (0.103), 0.01** | -0.07 (0.102), 0.493 | 0.03 (0.063), 0.634 |
| History of diabetes or medication | -0.035 (0.112), 0.758 | 0.023 (0.086), 0.793 | -0.052 (0.085), 0.543 | -0.012 (0.053), 0.819 |
| History of hypertension | 0.094 (0.089), 0.293 | **0.151 (0.069), 0.029** | 0.123 (0.068), 0.07 | -0.017 (0.042), 0.686 |
| Dementia in 5 years | 0.846 (0.12), <0.001 | 0.247 (0.093), 0.008 | 0.31 (0.091), 0.001 | -0.026 (0.057), 0.644 |
| Died in 5 years | 0.495 (0.117), <0.001 | 0.282 (0.091), 0.002 | 0.107 (0.089), 0.23 | -0.028 (0.056), 0.616 |

## Supplementary Material

### Comparison with RADAR

#### Quick Assessment of Dementia

The RADaR (Rapid Assessment of Dementia Risk) is a brief assessment tool designed to predict the risk of developing dementia in older adults by considering various factors like demographics, medical history, memory complaints, cognitive function, and functional abilities, allowing healthcare providers to identify individuals at high risk for further evaluation and intervention. The tool was designed for primary care settings and was validated in Black individuals. We calculated the RADaR using baseline age, memory complaint, the ability to handle finances, the recall of the month, recall of the room, and recall of three words as previously described^21^. The supplementary figure shows the time-specific, 3 to 7-year prediction, risk of dementia of pTau217, MMSE, and RADaR within the race. In this sample, RADaR had AUC_3 year_ of 0.80 (CI=0.70, 0.90) for Black and 0.78 (CI=0.72, 0.84) for White older adults, very equivalent to pTau217 alone. The discrimination of PTau217 and the RADaR was 0.85 (CI= 0.77, 0.94) for Black, and 0.80 (CI=0.733, 0.86) for White, therefore a small nominal gain compared to laboratory test alone, not enough to reach significance in this sample.

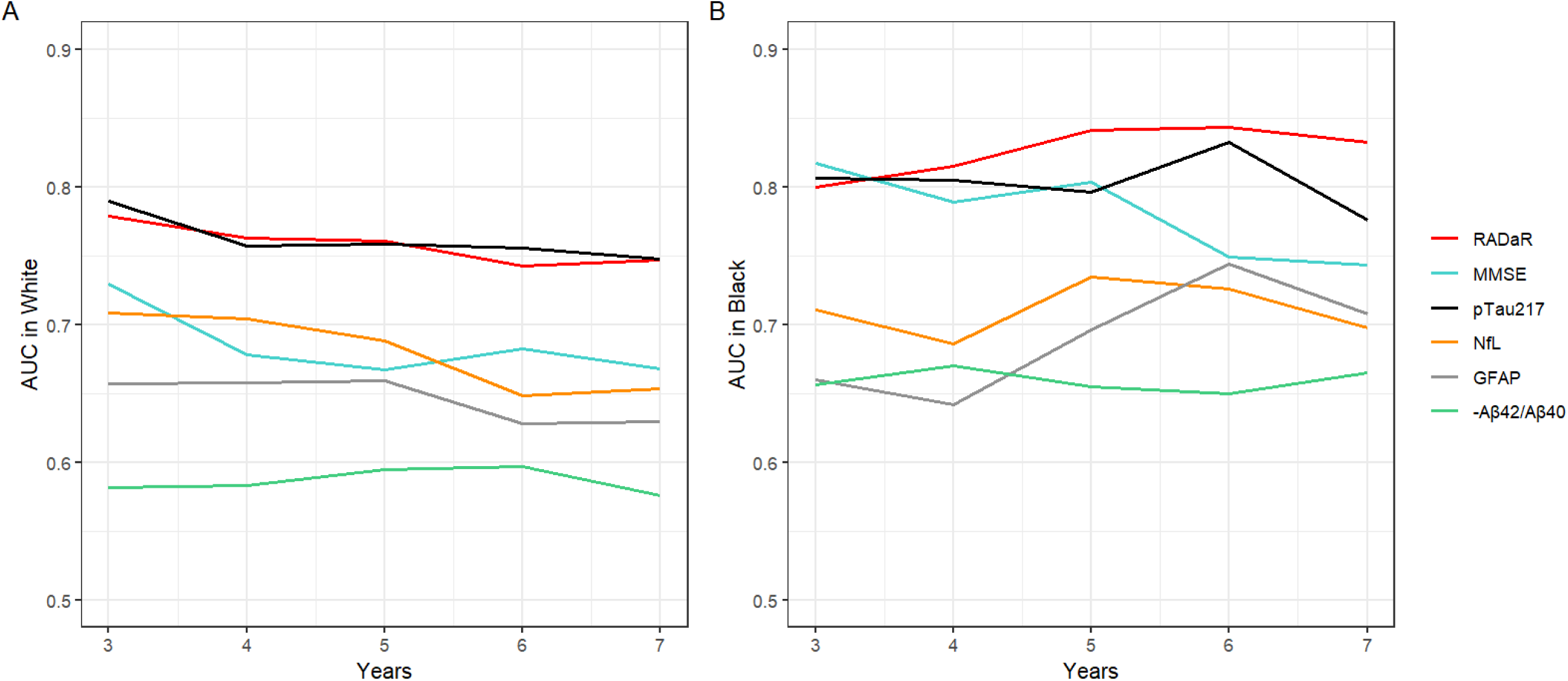
Supplementary Figure. Time-specific AUC of blood biomarkers for 3 to 7 years dementia discrimination. Graphs A and B include the total sample of White and Black older adults. GFAP =Glial Fibrillary Acidic Protein, NfL =Neurofilament Light Chain, Aβ42= beta-amyloid 1-42, Aβ40= beta-amyloid 1-40, and pTau217=phosphorylated tau at threonine 217.

## Appendices

### Study Level Biomarker Assay Quality Monitoring

The Biomarker Assay Laboratory (BAL) within the National Centralized Repository for Alzheimer’s Disease and Related Dementias (NCRAD) has implemented a quality program that includes bridging assays between instruments, assays, and reagent lots. The procedure is designed such that the laboratory reports consistent and reliable results over time and across studies. Briefly, a verification set of samples (n>25) is used to evaluate differences when major changes have occurred (e.g., assay, instrument or lot changes, instrument maintenance). If changes result in value differences outside of our normal operating criteria of 10% deviation, they are investigated, and bridging is performed based on the causes identified. This results in consistent and reliable return of results over time and across studies. In addition to the bridging procedure assay performance is monitored using three pooled plasma reference samples. These are included on each plate or within each batch depending on the instrument and batch design. The values are control charted and outliers are identified and investigated such that deviations outside of 20% across all three pooled plasma reference samples would necessitate failing a plate and not reporting those results. This monitoring ensures the quality of reported results and the ability to use prespecified cut points or reference ranges developed by the laboratory. If there are differences between studies, they will be a result of study-specific differences (e.g., study population, sample collection, or handling differences) and not attributable to laboratory operations.

The samples included in this study were run across many batches covering a total of 49 plates in two instances months apart. The average coefficient of variation across all 49 plates for each analyte is shown in Supplemental Table 1.

**sTable 1:** Study % coefficient of variation (%CV) determined for each of three pooled plasma reference samples. Results show all assays had ≤ 10% error across the two separate analyses and 49 plates.

| Assay | Pooled Plasma<br>Reference 1 Study %CV | Pooled Plasma<br>Reference 2 Study %CV | Pooled Plasma<br>Reference 3 Study %CV |
| --- | --- | --- | --- |
| N4PE - NfL | 7% | 6% | 8% |
| N4PE - GFAP | 7% | 5% | 6% |
| N4PE - Aβ1-40 | 4% | 3% | 4% |
| N4PE - Aβ1-42 | 5% | 5% | 5% |
| AlzPath P-tau217 | 8% | 10% | 10% |

